# Do professional fights go too long?: A video review of concussions and their consequences in professional boxing and MMA matches

**DOI:** 10.1101/2020.04.28.20073221

**Authors:** Charles Bernick, Tucker Hansen, Winnie Ng, Vernon Williams, Margaret Goodman, Bryce Nalepa, Guogen Shan, Tad Seifert

## Abstract

**Objectives:** Determine, through video reviews, how often concussions occur in combat sport matches, how well non-medical personnel can be trained to recognize concussions and how often fights are judged to continue too long.

**Methods:** This is a retrospective video analysis by an 8 person panel of 60 professional fights (30 boxing and 30 mixed martial arts). Through video review, medical and non-medical personnel recorded details about each probable concussion and determined if and when they would have stopped the fight compared to the official stoppage time.

**Results:** A concussion was recorded in 47/60 fights. The fighter that sustained the first concussion ultimately lost 98% of the time. The physician and non-physician raters had 86% agreement regarding the number of concussions that occurred to each fighter per match. The mean number of concussions per minute of fight time was 0.08 (0.06 for boxers and 0.10 for MMA). When stratifying by outcome of the bout, the mean number of concussion per minute for the winner was 0.01 compared to the loser at 0.15 concussions per minute. The physician raters judged that 24 of the 60 fights (11 boxing [37%]; 13 MMA [43 %]) should have been stopped sooner than what occurred.

**Conclusion:** Recognizing that the losing fighter almost always is concussed first and tends to sustain more concussions during the fight, along with the demonstration that non-physician personnel can be taught to recognize concussion, may guide policy changes that improve brain health in combat sports.

## Background

As our understanding of concussion and its’ potential lingering neurological effects has increased, there has been a heightened attention to concussion recognition in organized sports. While many contact sports have concussion protocols, where the injured athlete is pulled from competition until he is assessed to be back to his baseline neurological function, there is an inherent problem with this type of policy in combat sports (1). One of the implicit goals of a fighter is to induce a concussion on his/her opponent and render them unable to continue. Combat sport regulatory commissions vary on guidelines regarding concussions and who is authorized to stop a match. Generally, a contest is halted when a referee and/or physician decide an athlete can no longer defend him or herself (2). In many jurisdictions, the referee is the sole arbitrator authorized to end a fight while taking into consideration recommendations from the ringside physician (3). However, referees face immense pressure when making quick decisions to end a contest, especially during matches that can significantly influence the ranking or title of a combatant or their financial gain. During critical moments, referees are limited by visual perspective and their judgments can be indirectly influenced by crowd noise (4, 5), coaches’ exclamations (6), corner staff, promoters, and fans (both at the arena and the television audience). Yet, a fight stopped too late could lead to serious injuries and even death (7).

Few studies have examined the frequency and characteristics of concussions that occur during a sanctioned fight. Studies of video analysis in high contact sports have identified features that may indicate concussion including loss of consciousness, vacant stares, motor incoordination, and poor balance (8–10). Potentially, non-physician personnel could be trained to recognize these visible features of a concussion

The current study is aimed at determining how often concussions occur in combat sport matches, how well non-physician personnel can be trained to recognize concussions and how often would a physician stop the fight earlier than occurred. Ultimately, this type of data may help shape new concussion policies in combat sports.

### Methods

This was a retrospective video analysis study of 60 professional fights (30 boxing and 30 mixed martial arts [MMA]). The videos were reviewed by an 8 person panel consisting of 4 neurologists and 4 non-physicians familiar with combat sports (certified athletic trainer, college student, boxing inspector, combat sport promoter). Each panelist was instructed on the visual signs which would constitute a concussion. While we use the terminology, “concussion”, we realize that, technically speaking, it is a **suspected concussion**, since no direct examination of the fighter was performed. The reviewers watched each video on their own on a computer screen without the soundtrack to avoid bias by the commentators. Fight review data was recorded on a standard scoring sheet.

The 60 videos in the study were drawn from a consecutive series of professional televised fights beginning in May 2015. The 30 MMA fights reviewed were in the Ultimate Fighting Championship (UFC), and the 30 boxing fights reviewed were Premier Boxing Champions fights. To be included in the study, the fight must have been scheduled for a minimum of 6 rounds for boxing or a minimum of 3 rounds for MMA and have had a video publicly available of sufficient quality to reliably assess the behavior of the fighters. Among the 30 UFC fights selected, 11 were Lightweight fights (7 ending in TKO/KO; 4 ending by decision), 9 were Welterweight fights (6 ending in TKO/KO; 3 ending by decision), and 10 were Heavyweight fights (7 ending in TKO/KO; 3 ending by decision). Modified boxing weight classes were defined for the purposes of this study. Boxing fights with competitors less than 135 lbs were placed in the “Light” category, while those with competitors between 140 lbs–160 lbs were placed in the “Mid” category and those with competitors over 175lbs in the “Heavy” category. Among the 30 boxing fights selected, 10 were Light category fights (7 ending in TKO/KO; 3 ending by decision), 11 were Mid category fights (7 ending in TKO/KO; 4 ending by decision), and 9 were Heavy category fights (6 ending in TKO/KO; 3 ending by decision).

The set of 60 fights were divided into 4 unique subsets, each with 15 fights. Each subset included 7 or 8 boxing fights and 7 or 8 MMA fights sampled systematically. Each reviewer was given one of the 4 subsets of fights to review, such that exactly one physician and one non-physician reviewed each subset. The fight video reviewers recorded details about each observed concussion sustained by the combatants, including the round and time left in the round when the concussion occurred, the symptoms of concussion, the method of hit, and the location of the hit. The reviewer answered relevant questions about each fight, including whether or not they would have stopped the fight earlier than the official stoppage time, when they would have stopped it, and what the official stoppage time was..

The completed scorecards were sent by e-mail or mail to the Cleveland Clinic Center for Brain Health in Las Vegas Nevada and the data inputted for statistical analysis.

#### Statistical analysis

In the comparison of number of concussions, mean and standard deviation (SD) were reported for two fighting styles and the outcome of the bout. McNemar’s test was used to test the agreement between physician and non-physician on whether fights should be stopped sooner. For the agreement test, Kappa test was performed to test the agreement among rates. All these tests were two-sided at the alpha level of 0.05. All statistical analyses were performed by using SAS statistical software (Version 9.4; SAS Institute Inc., Cary, NC).

## Results

Of the 60 fights reviewed, a concussion was recorded as occurring by one of the physicians in 47 fights, and no concussion was recorded in the remaining 13 fights. The distribution of concussions fell preponderantly on the loser (Figure 1). While the winner of the fight generally did not sustain a concussion (or if so, rarely more than 1), the loser more frequently was allowed to sustain multiple concussions as depicted in figure 1. Moreover, in 46 of the 47 fights (98%) where a concussion occurred, the fighter that sustained the first concussion ultimately lost. The physician and non-physician raters agreed 86% of the time regarding the number of concussions that occurred to each fighter per match.

**Figure 1.**
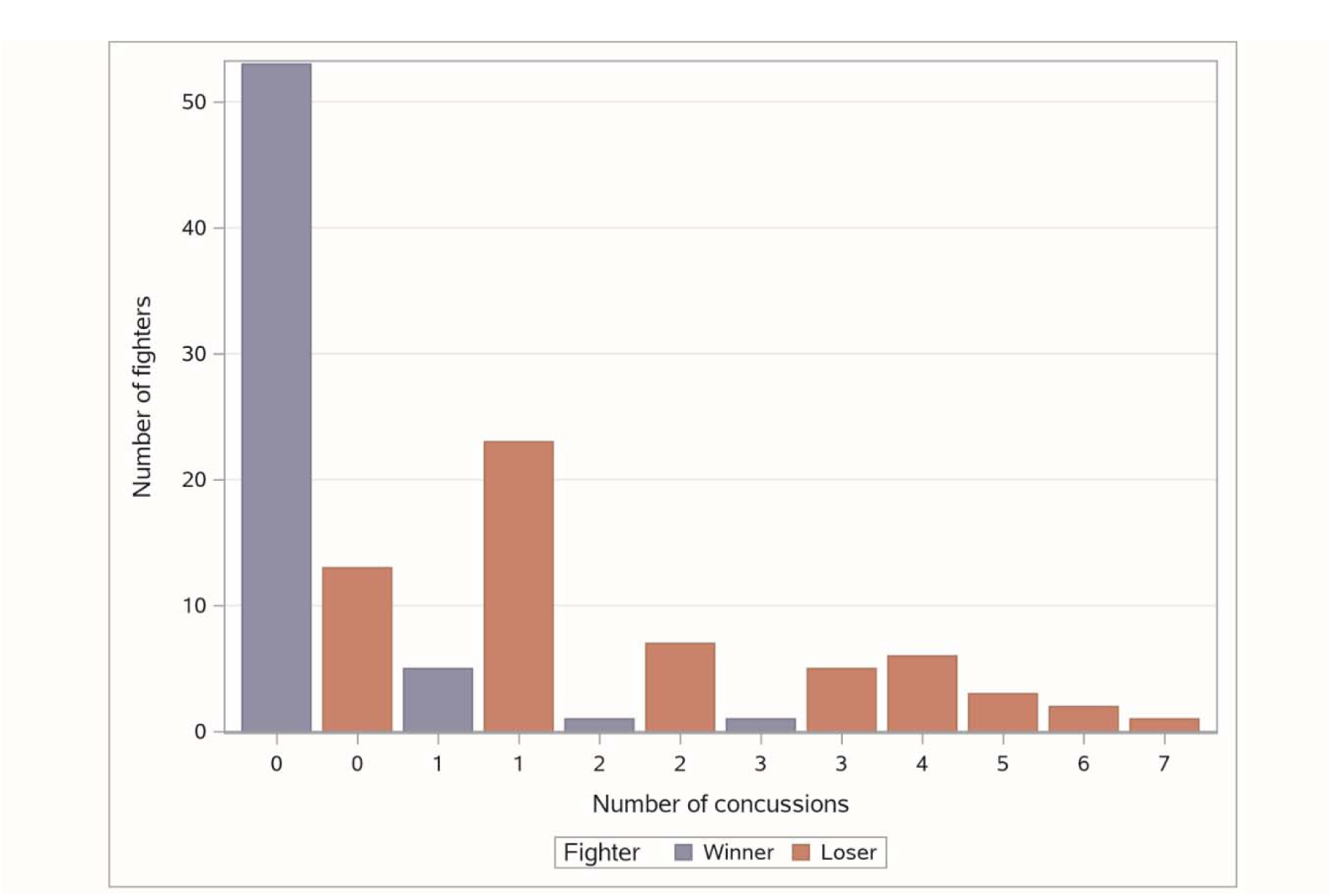
Number of concussions that were sustained by the winner and loser of each fight (n = 120 fighters: 60 winners and 60 losers)

Considering all the fights, the mean number of concussions per minute of fight time was 0.08 (0.06 for boxers and 0.10 for MMA). When stratifying by outcome of the bout, the mean number of concussion per minute for the winner was 0.01, whereas the loser on average sustained 0.15 concussions per minute (boxers only: 0.01/min for winner, 0.10/min for loser; MMA only: 0.01/min for winner, 0.20/ min for loser).

The panel of physician raters judged that 24 of the 60 fights (11 boxing [37%]; 13 MMA [43 %]) should have been stopped sooner than what occurred. The mean amount of time the fights were considered to go on too long was 125 seconds with a wide range extending from 3 to 835 seconds. Considering fighting disciplines, MMA fights seemed to terminate closer to what the reviewers had adjudicated, the mean time being 44.5 seconds (3-159 sec) compared to boxing matches that went on a mean 220 seconds too long (6-835 sec) (Figure 2).

**Figure 2.**
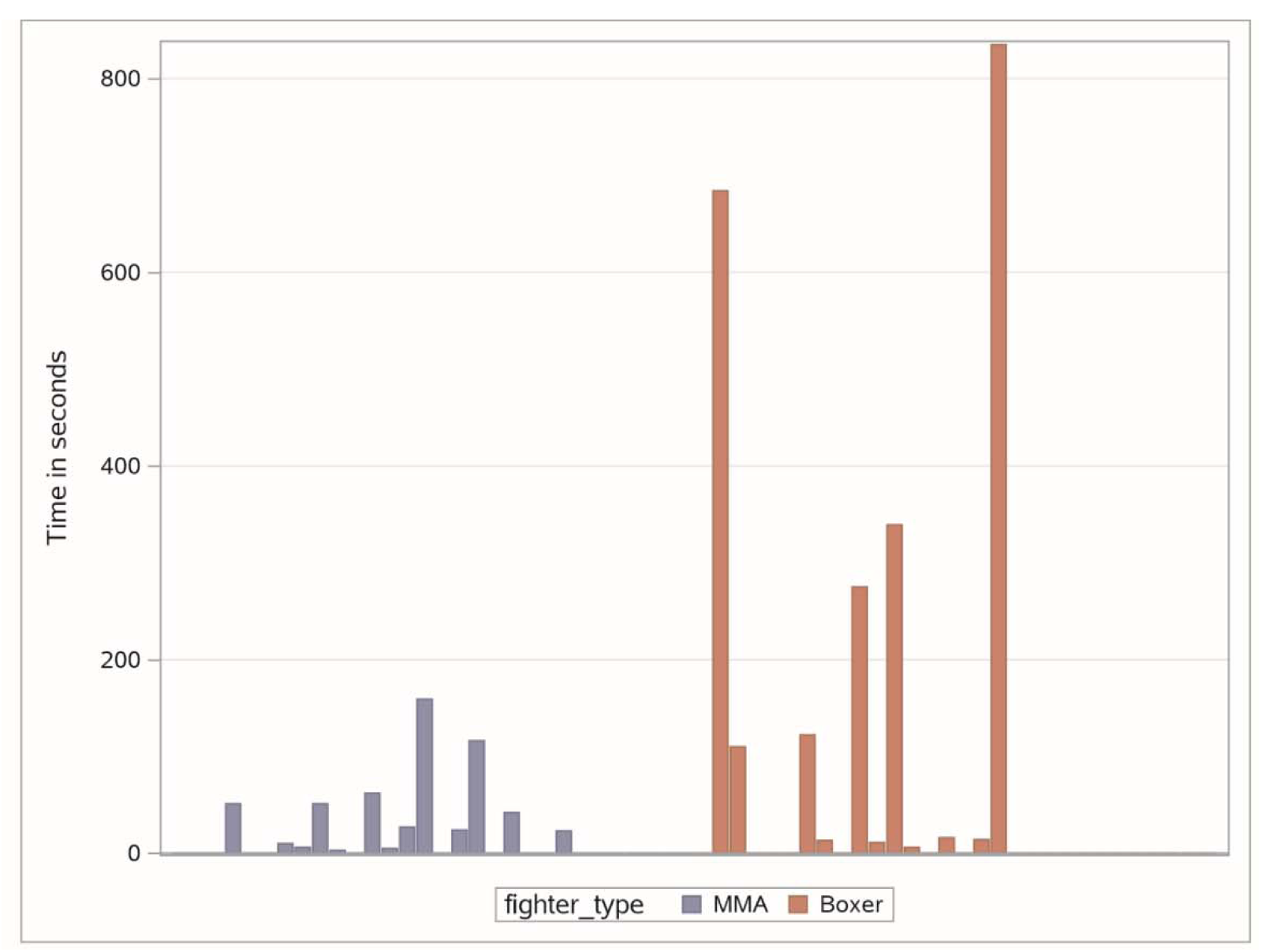
Time (in seconds) that a fight was considered by the neurologist reviewers to have continued too long. Each bar represents one fight.

In determining whether a fight should have been stopped sooner than it was, the non-physician raters agreed with the physicians’ judgment that the fight was stopped appropriately 85% of the time. On the other hand, the non-physicians only agreed on 63% of the fights that the physician thought went too long. The greatest discordance was with fights that the non-physician thought were stopped appropriately but the physician disagreed (8/50). The agreement index, Kappa coefficient, was 0.503, which is considered as moderate agreement between physician and non-physician (11), and the agreement test by using McNemar’s test had the p-value of 0.248.

Among the characteristics that identified a concussion, the fighter being knocked down was the most commonly cited symptom, followed by abnormal gait/balance, vacant stare/dazed, and delayed motor response, though the distribution of the later 3 varied slightly from boxers to MMA (Figure 3). While it was not always clear what specific blow was “causative” for the concussion, punches that landed on the right side of the face were most commonly linked to the concussion, with the temple being the next most common for MMA fighters and jaw for boxers.

**Figure 3.**
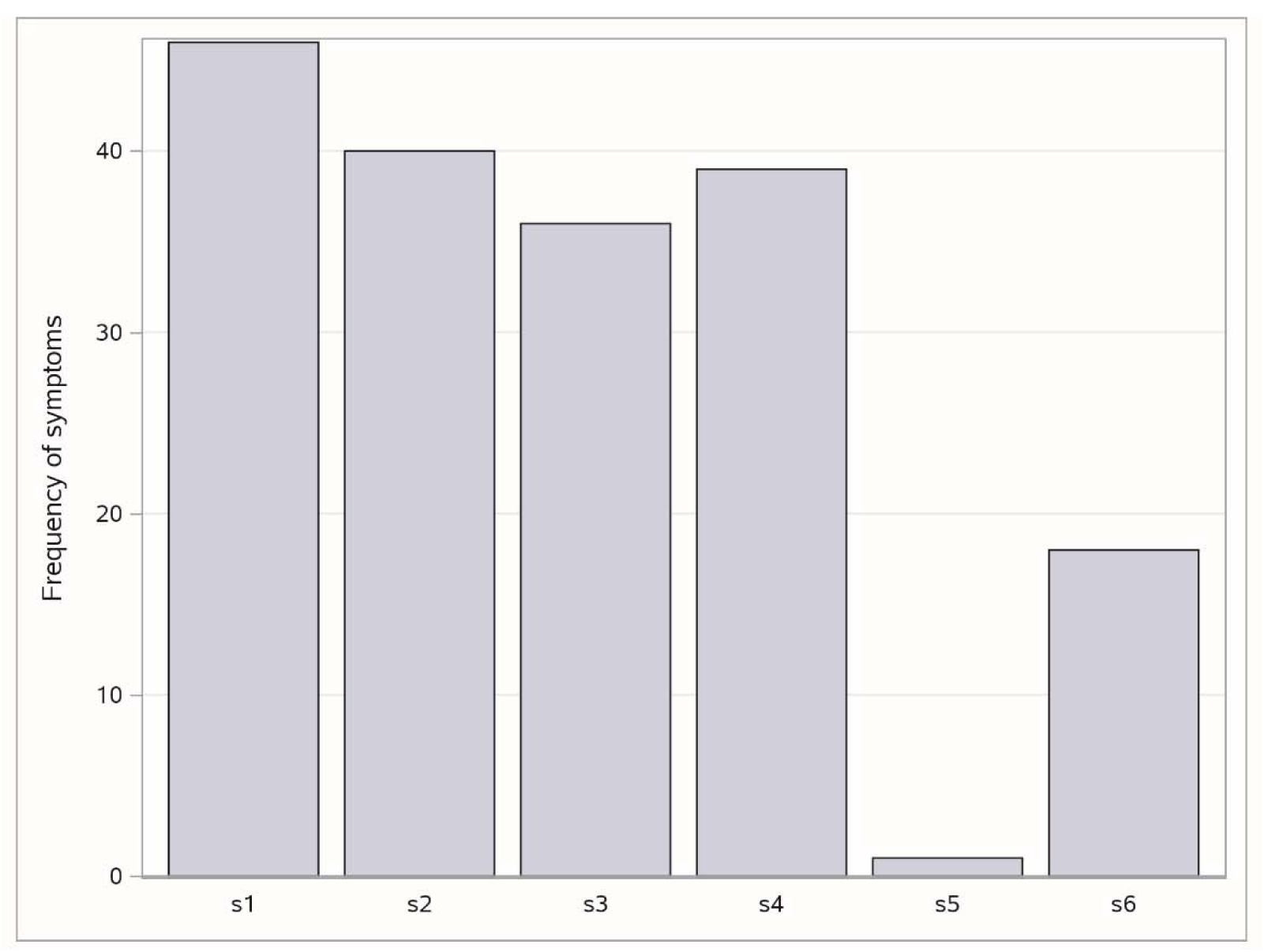
Number of times a specific visual sign was used to identify a concussion. A single concussion could have more than 1 sign. The signs area as follows: s1 Knocked Down, s2- Abnormal gait/balance, s3- vacant stare/dazed, s4- Delayed motor response (decreased ability to defend himself), s5- Focal neurological deficit, s6- Prolonged period of defensive positioning with no proactive attempts to strike opponent

## Discussion

Given the increasing attention to concussion and long term effects of exposure to repetitive head impacts, there is emerging interest in strategies to improve brain safety in sports. Historically, combat sports have been ripe for criticism since one of the major goals for competitors is to intentionally inflict neurologic trauma upon their opponents (12, 13). Yet, the majority of fights do not end in knock outs and there may be policy changes that can make combat sports safer in the 21^st^ century (1, 14). Along those lines, this study was aimed at better understanding the frequency of observed concussions in sanctioned competitions, how their timing effects the outcome of the fight, and how often fights are judged to go on too long.

### Frequency of concussion

Among the most notable findings was that recognized concussions are likely to occur in a fight, averaging about 1 every 12.5 minutes of fighting time. What perhaps was most striking in our series was that the fighter that sustained the first concussion in a bout/match almost always went on to lose. Moreover, many losing fighters sustain multiple concussions in a fight. These findings, along with the judgement by neurologist reviewers that close to 40% of the fights should have been stopped sooner, have several practical implications.

Unlike the current status in many professional sports leagues, where an athlete suspected to have sustained a concussion are removed from the game, that is not necessarily the case in combat sports (15–17). It becomes the judgment of the referee (at times with input from the ringside physician) as to whether a fighter should continue despite experiencing a concussion. Referees and ring physicians thus must balance the risk of stopping a fight prematurely denying the concussed fighter the chance of possibly coming back and winning (which may have significant financial implications) versus letting that fighter unnecessarily absorb further brain injury. And although, many experienced referees are quite good at assessing the status of fighters, there are a number of external pressures that may exert some influence to allow the fight to go on as long as possible including the stature of the match (championship v. undercard), expectations from paying event attendees, television and pay per view viewers, gaming interests, promoters and the fighters themselves.

Fighters that display concussion signs, yet rapidly normalize, are often allowed to continue. Whether it is because having experienced a concussion makes one more susceptible to another, impairs ability to compete as well, or that the inferior fighter is most likely to be concussed first, the fighter that sustained the first concussion in our video series were overwhelmingly *unlikely* to ultimately win. This finding may be surprising to those who are veteran observers of combat sports, as there are certainly plenty of examples of a fighter who seems to absorb a major hit first but then comes back to win. While our sample size is relatively low (60 fights), we did have a consecutive series of high level, relatively well matched fights which lends some credence to our result.

### Do fights go on too long

Given the luxury of watching the replay of a fight, our reviewers felt a significant proportion of fights were allowed to continue past the point where it seemed visibly clear that one of the fighters was impaired. The amount of time between when the physician reviewer would have stopped the fight and the official stop time tended to be shorter for MMA compared to boxing matches. This finding is likely due to the different rules governing the two sports. If an MMA fighter is knocked to the ground, the fight still continues on and the fighter has no reprieve to recover; the opponent can continue to strike the fighter with the referee closely observing (18). In boxing, once a fighter is knocked down, he/she has 8 protected seconds to recover and simply needs to show the referee that they can continue.

### Concussion recognition

While there is an entire industry based on devising techniques and devices to detect concussion, our study suggests that non-physician personnel can be trained to reliably recognize the visual signs of concussion (19). In 86% of cases, both the physician and non-physician recorded the same number of concussions in a match. When there was disagreement, more often the non-physician assessed a concussion while the physician didn’t. It is possible the non-physicians felt a heightened vigilance to not miss a concussion. From this data, it would seem that, even though there is a physician at ringside during sanctioned matches, it is certainly feasible that all other officials at ringside including referees, judges, and inspectors can be trained in concussion recognition. In addition, in fights where there is only one ringside physician, certified athletic trainers (ATC) who are already familiar with concussion protocols could provide additional surveillance during the match and inspectors could be responsible for observing behavior of fighters in their corner between rounds and alerting the physician or referee if any signs of concussion are noted.

### Concussion protocol in combat sports?

How our findings and others can be translated into policies that improve safety in combat sports is challenging. Many of the major promoters already have put into place standards that exceed the minimum requirements by the state regulatory commissions. For example, the UFC requires 4 ringside physicians, along with having their own medical director present at ringside who monitor the fighters during and in-between rounds (Davidson J, personal communication). However, other promotions may have only one physician present, which reduces the ability to scrutinize the ongoing status of both fighters. Furthermore, a referee may be hesitant to stop a fight in mid-round for suspected concussion (particularly early in the fight) unless the fighter is clearly impaired.

Yet, one could envision implementing a concussion policy in combat sports. This may consist of a mandatory assessment of a fighter between rounds if the referee, physician, or ATC witnesses a concussion or if the inspector has concerns about the fighter between rounds. The assessment could consist of a brief standardized assessment in the corner by the fight doctor; the fight would be stopped if the fighter demonstrated amnestic or psychomotor deficits. In addition, the sport of boxing could decrease the standing 8 count to 4 seconds, which would allow fighters who fell to the ground through a slip or body punch or who are able to rapidly clear their overt signs and symptoms of concussion to continue, yet protect those who sustain a greater degree of motor or cognitive damage. One might argue that if concussions occur on average every 12.5 minutes, then with these policies most boxing matches would end within 4 to 5 rounds and MMA within 3 rounds. However, a number of fights reviewed had no observed concussions including championship fights that went the distance. Moreover, some fighters do not immediately show signs of impairment after a suspected concussion and continue to compete despite their unrecognized injury. Nevertheless, the fighter that sustained the first concussion should always have particular scrutiny throughout the match.

Even in the absence of a concussion protocol in combat sports, at the minimum, everyone involved in the fighters’ safety at ringside should undergo concussion training. Due to the competitive nature of combat sports and the unlikelihood of fighters voluntarily quitting or stopping a fight, it is critical that referees, physicians and other ringside personnel work cohesively and are well trained to identify serious injuries in order to stop fights in a timely manner (7). Having regulatory commissions that sanction fights require specific concussion training to referees and other ringside personnel would be an important safety measure. Criteria for fight stoppage should be standardized across all combat sports, in addition to in-fight injury assessment. Despite potential conflicts of interest due to a desire for competitive success, the corner staffs’ primary responsibility is to protect the safety of their associated fighter. Requirement for licensing of trainers should also include concussion recognition.

Admittedly, it is not known if addressing concussions in combat sports and ending fights sooner would benefit a fighter’s overall brain health. A significant amount of the long term neurological damage has been reported to occur with routine sparring during training (20, 21). However, there are anecdotal cases of fighters who seemingly were allowed to sustain cumulative blows to the head despite overt signs of neurologic impairment; thus resulting in clear neurological morbidity (22). Our study also has a number of obvious limitations including a relatively small number of fights reviewed. All bouts reviewed were high level fights and may not be generalizable to lower level fights in smaller venues. Lastly, without actually examining the fighter at the time or being able to review post-fight medical records, we cannot be sure what we assess as a concussion was genuinely a concussion.

Recognizing that the losing fighter almost always is the one that is concussed first and tends to subsequently sustain more concussions during the fight, along with the demonstration that non-physician personnel can be taught to recognize concussion, can hopefully help guide policy changes that improve brain health in combat sports.

## Data Availability

data is available upon reasonable request

## Acknowledgement

The authors would like to recognize the contributions made by Mr. Todd Neal and Mr. Gary Thomas in providing their expertise in combat sports through video reviews.

## Notes

### Competing Interest Statement

Charles Bernick has received research support from the UFC, Haymon Boxing and Top Rank Promotions

### Funding Statement

No external funding was received for this manuscript

## References

1. Seifert T, Bernick C, Jordan B, et al. Determining brain fitness to fight: Has the time come? Phys Sportsmed. 2015 Nov; 43(4):395–402. doi: 10.1080/00913847.2015.1081551

2. Miele, V. J., & Bailes, J. E.. Objectifying when to halt a boxing match: A video analysis of fatalities. Neurosurgery. 2007; 60(2), 307–316. doi:10.1227/01.NEU.0000249247.48299.5B.

3. Kelly, M. Role of the ringside physician and medical preparticipation evaluation of boxers. Clinics in Sports Medicine. 2009; 28(4), 519, v. doi:10.1016/j.csm.2009.07.003.

4. Balmer, N, Nevill, A, Lane, A, et al. Influence of crowd noise on soccer refereeing consistency in soccer. J. Sport Behav. 2007; 30(2), 130–145.

5. Nevill, A. M., Balmer, N. J., Williams, A. M. The influence of crowd noise and experience upon refereeing decisions in football. Psychology of Sport and Exercise. 2002; 3(4), 261–272. doi:10.1016/S1469-0292(01)00033-4.

6. Souchon, N., Fontayne, P., Livingstone, A et al. External influences on referees’ decisions in judo: The effects of coaches’ exclamations during throw situations. Journal of Applied Sport Psychology. 2013; 25(2), 223–233. doi:10.1080/10413200.2012.713440.

7. Sethi, N. K. Good versus bad medical stoppages in boxing: Stopping a fight in time. South African Journal of Sports Medicine. 2016; 28(3), 87–89. doi:10.17159/2078-516x/2016/v28i3a1735

8. Bruce, J. M., Echemendia, R. J., Meeuwisse, W et al. Development of a risk prediction model among professional hockey players with visible signs of concussion. British Journal of Sports Medicine. 2018; 52(17), 1143–1148. doi:10.1136/bjsports-2016-097091.

9. Davis, G., Makdissi, M. Use of video to facilitate sideline concussion diagnosis and management decision-making. Journal of Science and Medicine in Sport. 2016; 19(11), 898–902. doi:10.1016/j.jsams.2016.02.005.

10. Gardner, A., Howell, D., & Iverson, G. A video review of multiple concussion signs in National rugby league match play. Sports Medicine-Open. 2018; 4(1), 1–8. Doi: 10.1186/s40798-017-9

11. Shan, G, Wilding, G.E.,. Powerful exact unconditional tests for agreement between two raters with binary endpoints. PloS one, 2014; 9(5), p.e97386.

12. American Academy of Pediatrics, Council on Sports Medicine And Fitness; Canadian Paediatric Society, Healthy Active Living And Sports Medicine Committee, Purcell L, LeBlanc CM. Policy statement—Boxing participation by children and adolescents. Pediatrics. 2011 Sep;128(3):617–23. doi: 10.1542/peds.2011-1165.

13. Sammons JT. Why physicians should oppose boxing: an interdisciplinary history perspective. JAMA. 1989 Mar 10;261(10):1484–6.

14. Bernick C, Banks SJ, Shin W, et al. Repeated head trauma is associated with smaller thalamic volumes and slower processing speed: the Professional Fighters’ Brain Health Study. Br J Sports Med. 2015 Aug;49(15):1007–11. doi: 10.1136/bjsports-2014-093877

15. Concussion policies by league. USA Today. 2012. Available from http://www.usatoday.com/story/sports/2012/10/11/concussions-nascar-nfl-mlb-nhl-nba/1628129/. Last accessed 5 November 2014.

16. National Basketball Association. Concussion policy summary. Available from http://www.nba.com/official/concussion_policy_summary.html. Last accessed 5 November 2014.

17. Major League Baseball MLB. Union adopt universal concussion policy. Available from http://m.mlb.com/news/article/17183370/. Last accessed 5 November 2014.

18. Stephen SJ, Shan G, Banks SJ, et al. The Relationship Between Fighting Style, Cognition, and Regional Brain Volume in Professional Combatants: A Preliminary Examination Using Brief Neurocognitive Measures. J Head Trauma Rehabil. 2019 Nov 8. doi: 10.1097/HTR.0000000000000540.

19. Okonkwo DO, Tempel ZJ, Maroon J. Sideline assessment tools for the evaluation of concussion in athletes: a review. Neurosurgery. 2014 Oct;75 Suppl 4:S82–95. doi: 10.1227/NEU.0000000000000493.es

20. Bernick C, Zetterberg H, Shan G, et al. Longitudinal Performance of Plasma Neurofilament Light and Tau in Professional Fighters: The Professional Fighters Brain Health Study. J Neurotrauma. 2018;35(20):2351–2356. doi: 10.1089/neu.2017.5553

21. Stiller JW, Yu SS, Brenner LA, et al. Sparring and neurological function in professional boxers. Front Public Health. 2014;2:69. doi: 10.3389/fpubh.2014.00069

22. Park A. The role of boxing in the death of Muhammad Ali remains unclear. Time. 2016 Jun 20;187(23):12.

